# Opt-out policies capacity to increase organ donors is limited

**DOI:** 10.1101/2021.08.27.21262033

**Authors:** Alberto Molina-Pérez, David Rodríguez-Arias, Janet Delgado

## Abstract

**Objectives:** To increase post-mortem organ donation rates, several countries are adopting an opt-out (presumed consent) policy, meaning that individuals are deemed donors unless they expressly refused so. However, studies on the relative impact of opt-in or opt-out on deceased organ donation rates are inconclusive. Although opt-out countries tend to have higher donation rates, there is no conclusive evidence that this is caused by the policy itself. The main objective of this study is to better assess the impact of consent policies when considering the role of the family in decision-making.

**Design:** By systematically combining the three components of the decision-making process —the default rule, the deceased’s preferences, and the family’s preferences,— we identify all situations that affect the retrieval outcome under opt-in and opt-out policies. Then, by gathering empirical data from a wide array of countries, we estimate the relative frequency of these situations.

**Main outcome measures:** We measure the relative impact that opt-in and opt-out policies have *per se* on post-mortem organ retrieval.

**Results:** Our analysis shows that opt-in and opt-out have strictly identical outcomes in eight out of nine situations. These policies only differ when neither the deceased nor the family have expressed a preference and defaults therefore apply. The actual impact of consent policies is typically circumscribed to a range of 0% to 5% of all opportunities for organ retrieval.

**Conclusions:** This study may warn contemporary organ retrieval policy makers that, by emphasizing the need to introduce presumed consent, they might be overestimating the influence of policy defaults and underestimating the power granted to families in expressing their preferences and making decisions about organ donation. Governments should reassess the opportunity and effectiveness of adopting opt-out policies for organ retrieval.

What is already known on this topic

- Studies on the relative impact of opt-in and opt-out on deceased organ donation rates are inconclusive.
- There is a correlation between presumed consent and higher rates of organ retrieval, but no evidence of a causal relationship.
- Most studies overlook the role of the family in decision-making.

What this study adds

- When the role of the family is taken into account, opt-in and opt-out policies have identical outcomes in eight out of nine situations.
- The situation where opt-in and opt-out actually differ from each other typically occurs in less than 5% of post-mortem organ retrieval opportunities.
- Moving from opt-in to opt-out can only marginally increase the organ supply.

Strengths and limitations of this study

- The main strength of this study is the combination of analytical and empirical methods.
- This is the first study to analyse all situations that affect the retrieval outcome under both opt-in and opt-out policies when considering the role of the family in decision making.
- Data analysed in this study is the best empirical evidence available to date.
- The main limitation of our study is the heterogeneity of sources, sample sizes and time periods for the data collected, especially for the additional supporting evidence.
- This study only considers the direct effects of opt-out policies on organ retrieval rates, but not its indirect effects, such as organ preservation measures and psychological effects.

## Introduction

There is an international trend to move from explicit consent (opt-in) to presumed consent (opt-out) policies for deceased organ retrieval: Chile (2010), Finland (2010), Greece (2013), Uruguay (2013), Wales (2015), Colombia (2016), Iceland (2019), the Netherlands (2020), England (2020), Scotland (2021), and the province of Nova Scotia in Canada (2021) have implemented opt-out policies. Switzerland is presently considering it, and Australia, Denmark, Germany, Israel, Romania, and several states in the USA had been discussing this as well.

Policy changes towards opt-out seek to increase donation rates by adding to the pool of potential donors all deceased adults who did not express an objection while alive. Some studies suggest that presumed consent laws indeed contribute to increased organ donor rates[1–6], while others dispute this claim[7–11]. Research reviews within this field point out an association between presumed consent legislation and increased organ donation rates, but they also warn against the assumption that the introduction of presumed consent legislation *per se* leads to an increase in organ donation rates[12–14]. Consent policies may, in fact, be just one factor among many, with infrastructure or organisational changes producing greater gains than legislative change alone[15,16]. The role families are allowed to play in the process of organ retrieval decision-making may be another factor tempering the effectiveness of presumed consent policies[8,17–19]. While most studies on this subject have overlooked the role of the family, the interaction between consent systems, deceased’s decisions, and families’ preferences deserves further clarification.

In this article, we examine how such a relationship impacts the outcome of organ retrieval decision-making (i.e. whether it enables or hinders organ recovery). To do so, we developed a novel approach. First and foremost, we provide analytical evidence of the differential impact that opt-in and opt-out policies can have *per se* on organ retrieval rates, that is, regardless of the country they are implemented in. Additionally, we provide confirmatory evidence for these analytical results based on the best empirical data available, that is, comprehensive high quality data from six European nations and partial data from 15 other countries worldwide. Finally, we estimate how changing to a different policy would affect the potential retrieval rates in seven countries.

## Methods

The development of the research question and outcome measure was informed by the results of a systematic review on public knowledge and attitudes towards consent policies for organ donation[20] and by a conceptual framework of the role of family in organ retrieval decision-making[21]. The review’s results suggested, on the one hand, that people’s awareness of the consent model is lower in opt-out countries than in opt-in countries, which raises ethical concerns with regard to the respect of individual autonomy, and, on the other hand, that despite the general tendency in Europe and elsewhere to move from opt-in to opt-out policies, a majority of the public tend to prefer opt-in and mandatory choice to opt-out when two or more options are offered. The framework’s results suggested that there is no significant difference between opt-in and opt-out policies when family preferences are considered.

### Analytical approach

We used the following analytical approach to assess how consent policies can impact organ retrieval rates. This approach allows for an examination of the consent policies *per se*, regardless of country-specific confounding factors such as organisation and infrastructures, professionals’ training, incentives, media campaigns, cultural backgrounds, etc.

First, we broke down consent policies into their core components[22]. As their name suggests, opt-in and opt-out policies are relative to individual preferences. This is the first element to consider. Organs may be retrieved when people expressed their consent as *post-mortem* organ donors (opt-in) and they may not be retrieved when people expressed their refusal (opt-out). In some countries, such as Germany and the Netherlands, individuals can also chose to delegate the decision to their relatives or a designated proxy. This introduces family preferences as a second element to consider. Indeed, whether the deceased’s organs are recovered or not may eventually depend on the next-of-kin’s attitudes towards donation. The third element is the default option set by each policy when no preferences have been expressed whatsoever. In such circumstances, organs can nevertheless be retrieved under opt-out, based on presumed consent, whilst they cannot be retrieved under opt-in.

The procedures deemed valid to express a preference regarding organ donation are also an important part of consent policies. These procedures may include consent and/or refusal registries, organ donor cards, living wills and other written documents, as well as conversations with relatives. Although some of these procedures can exist in a given country, they may be inconsequential as long as people are unfamiliar with them. For example, in France, the refusal register is by law the main procedure to express a decision, but less than 0.5% of the total population were listed in it by 2017[23]. For the sake of simplicity, considering the diversity and varying degrees of use of these procedures, we decided not to include them in our analysis. In the following analysis, we will consider the preferences of individuals and relatives, and the role they play under each policy, regardless of the means by which these preferences can be expressed in any given country.

Secondly, based on the aforementioned core components of opt-in and opt-out (individual preferences, family preferences, and defaults), we identified all the situations where the retrieval outcome depends on individual and/or family preferences or the lack thereof[22]. When relatives’ preferences are *not* taken into account, only three possible situations arise, as the deceased person may have either: (*A*) expressed their consent to donate; (*B*) expressed their refusal to donate; or (*C*) failed to express any decision regarding donation, in which case the default applies. When relatives *are* consulted, their own preferences regarding the recovery of organs from their loved ones may be either (*a*) favourable, (*b*) unfavourable, or (*c*) unknown. The combination of the preferences of the deceased and those of the family thus creates a total of nine (3×3=9) situations (Table 1).

**Table 1.** Consent-related situations that affect the retrieval outcome under both opt-in and opt-out policies.

|  |  | Family preferences |  |  |
| --- | --- | --- | --- | --- |
|  |  | a) Favourable | b) Unfavourable | c) Unknown |
| Deceased's preferences | A) Consent | Agreement in favour | Conflicting preferences | Deceased's consent |
|  | B) Refusal | Conflicting preferences | Agreement against | Deceased's refusal |
|  | C) Unknown | Family authorisation | Family opposition | Default applies |

Thirdly, we compared the outcomes of opt-in and opt-out policies in these nine situations. For example, whenever the deceased expressed their willingness to donate (*A*) and the family also expressed their authorisation (*a*), organ recovery is most likely to proceed in either opt-in and opt-out policies. This way, by comparing the two policies in each and every scenario, the identification of the situations producing the same retrieval outcome under both policies and those producing variable outcomes is straightforward. This allowed us to evaluate the relative impact of opt-in and opt-out policies *per se*, regardless of the country-specific confounding factors where these policies are implemented.

### Review of the empirical data available

#### Search strategy and data collection

To check whether our analytical findings were consistent with real-world national figures, we sought empirical data on the expression of preferences or the lack thereof, from either the deceased person or their relatives, in all cases of potential or eligible deceased donors. We included articles (scientific studies) and statistics from governments and transplant organisations (grey literature) from the last ten to fifteen years. We searched on Pubmed, Google Scholar, and ResearchGate using the following keywords in three languages (English, Spanish, and French): potential donor(s), potential organ donor(s), organ donor audit, potential organ donation, organ donation activity, organ donation referral, organ donation statistics, organ transplantation statistics, and the MeSH terms: “Tissue and Organ Procurement/statistics and numerical data” and “Organ Transplantation/statistics and numerical data”. We also searched directly into the websites of several national transplant organisations, when available. Additionally, we contacted national officials and researchers to help us locate relevant data, if any. Our search specifically targeted—but was not restricted to—55 countries from the five continents with deceased organ donation programmes.

#### Data extraction and quality assessment

All data used in this study were publicly available prior to the initiation of this study. Relevant data was extracted from two main sources of information: official national reports on the one hand (grey literature), and local retrospective studies on the other (scientific articles). To ensure accuracy, we contacted representatives of health ministries and national transplant organisations for clarifications or for confirmation of our findings. Evidence for the Netherlands, the United Kingdom, and Denmark have been double-checked and confirmed through personal communications with the *Nederlandse Transplantatie Stichting* (NTS), the National Health Service (NHS), and the *Dansk Center for Organdonation*, respectively. Evidence for Germany have been checked with the assistance of German researchers on organ donation. In a few instances, when no written source of information was available, we contacted the heads of national transplant organisations and other officials for comments. More detailed information about the sources and methods is available in a Supplementary File.

### Patient and Public Involvement

No patient involved.

## Results

### Analytical evidence

The three core components of consent policies that influence the outcome of the decision-making process (organ retrieval or non-retrieval) are: (i) the deceased’s expressed preferences, if any; (ii) the next-of-kin’s preferences, if any; and (iii) the default option set by each policy.

When family preferences are *not* taken into account, a side-by-side comparison of opt-in and opt-out policies shows that they have identical retrieval outcomes in two out of three situations, that is, whenever the deceased had either consented or refused organ donation. These two policies only differ in one situation: when the deceased person failed to express any decision and the default therefore applies (Table 2). In this situation, the absence of an explicit consent precludes organ retrieval under opt-in while the absence of an explicit refusal allows it under opt-out.

**Table 2.**
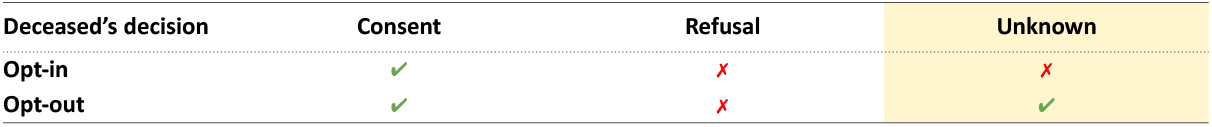
Outcome (organ retrieval vs non-retrieval) from organ recovery decision-making based on the deceased’s decision and the model of consent.

When *both* the individual and the family preferences are taken into account, a side-by-side comparison of opt-in and opt-out policies shows these policies having rigorously identical outcomes in eight situations out of nine. The lone situation when these policies make a difference is when their defaults apply, that is, when the preferences of both the deceased and their family remain unknown to the medical team (Table 3).

**Table 3.**
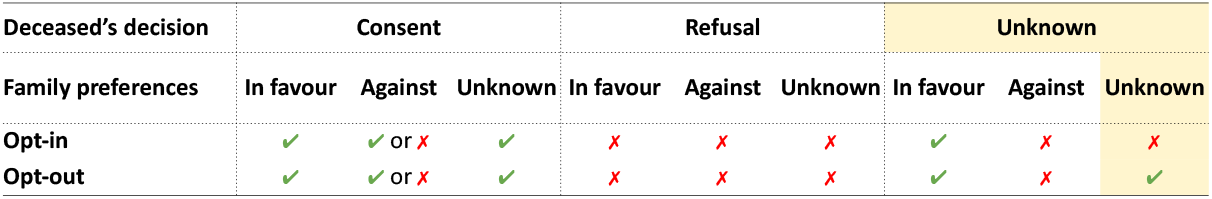
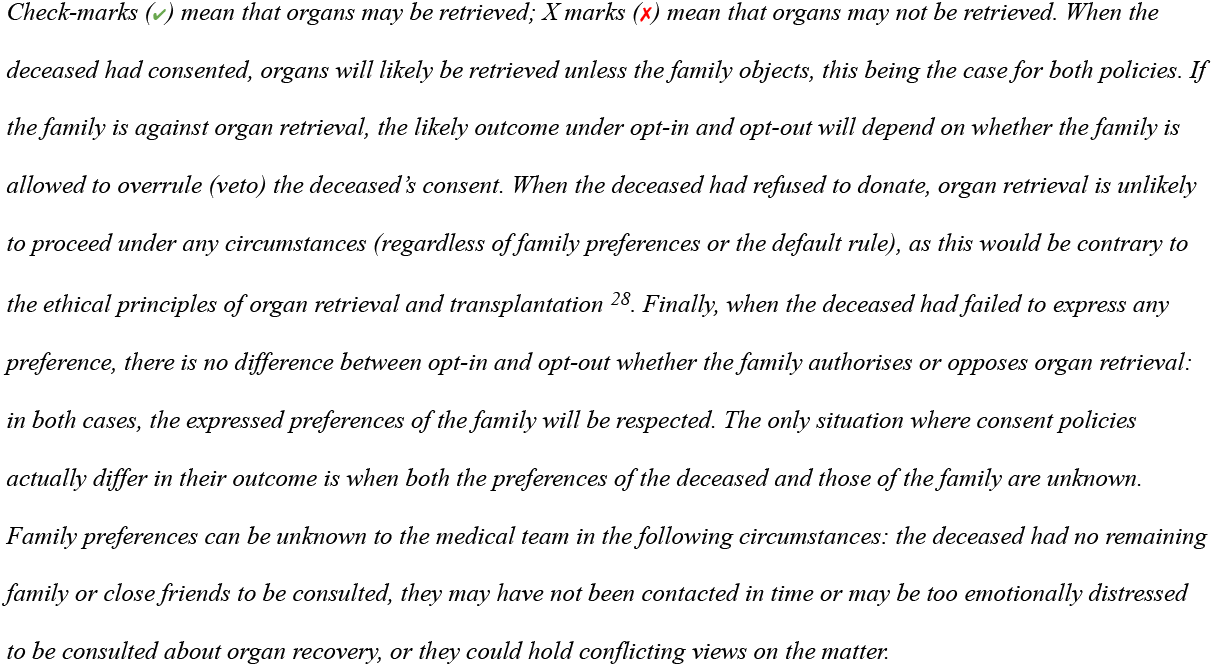
Outcome (organ retrieval vs non-retrieval) from organ recovery decision-making based on the deceased’s decision, family attitudes, and the model of consent.

This analysis shows that the differential impact of opt-in and opt-out policies is entirely determined by the default, which applies only when preferences have *not* been expressed. If all opportunities for organ retrieval were evenly distributed, the default would apply in one out of three cases (33%) in countries where relatives are not consulted (cf. Table 2), and one out of nine cases (11%) in countries where relatives are consulted (cf. Table 3).

### Empirical evidence

We obtained relevant empirical data from 21 countries in the five continents. Considering the diversity of sources and varying quality of the data, we classified the evidence obtained in two tiers: confirmatory evidence and additional supporting evidence. *Confirmatory evidence* includes comprehensive statistics from either government backed official reports or high-quality retrospective studies. *Additional supporting evidence* includes partial statistics from official reports and retrospective studies.

#### Confirmatory evidence

We found comprehensive nationwide statistics from official sources in Germany, the Netherlands, and the United Kingdom, and from peer reviewed retrospective studies in Sweden and Wales. In addition, we found comprehensive statistics from a retrospective study of all patients who died at one of the largest hospitals in Denmark between 2000-2003 and 2007-2010. Our findings show that, when families intervene, the differential impact of opt-in and opt-out policies, based on actual empirical evidence from these six countries, is limited to a range of 0% to 5% of all organ retrieval opportunities (Table 4; see supplementary file for more detailed information about the data, sources, and methodology).

**Table 4.**
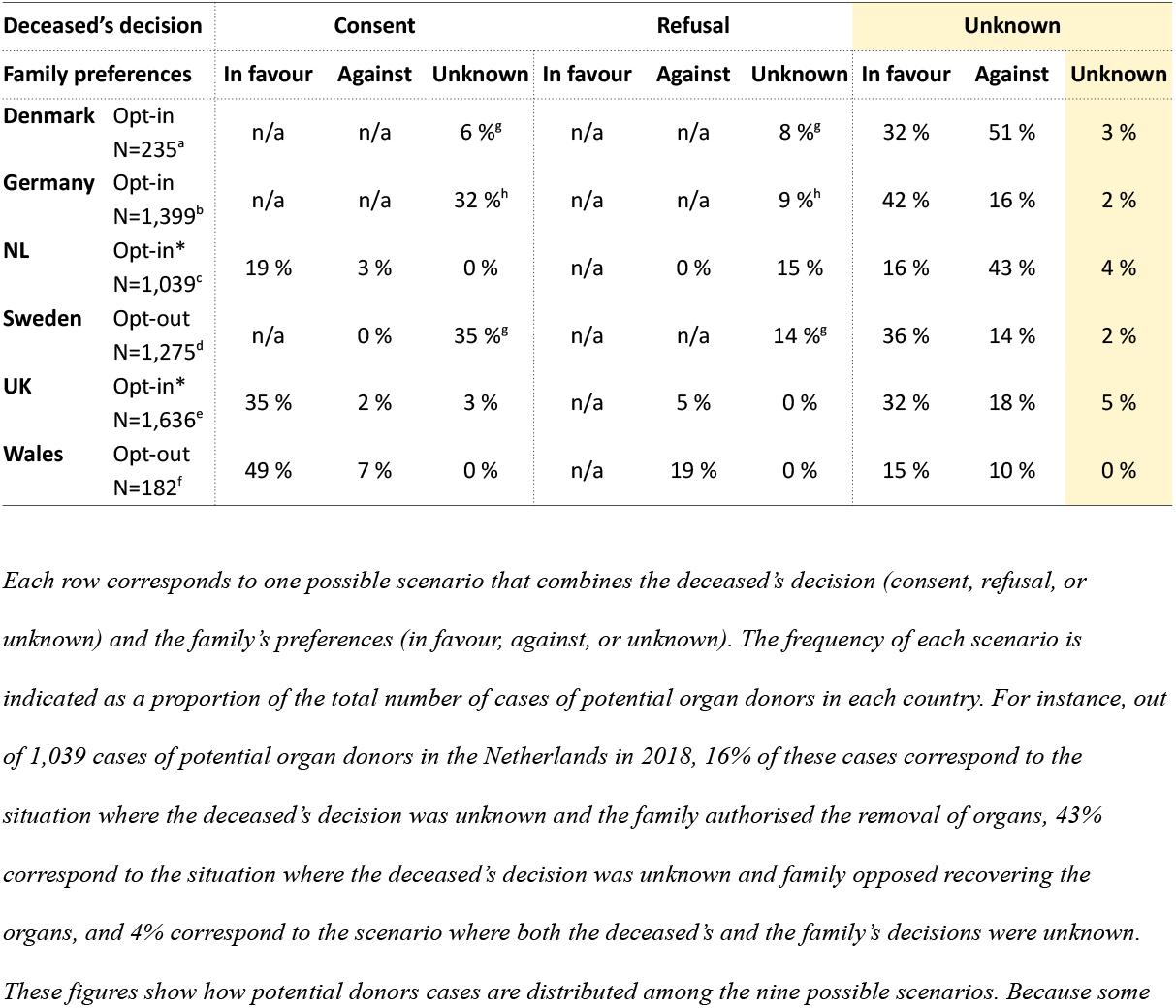
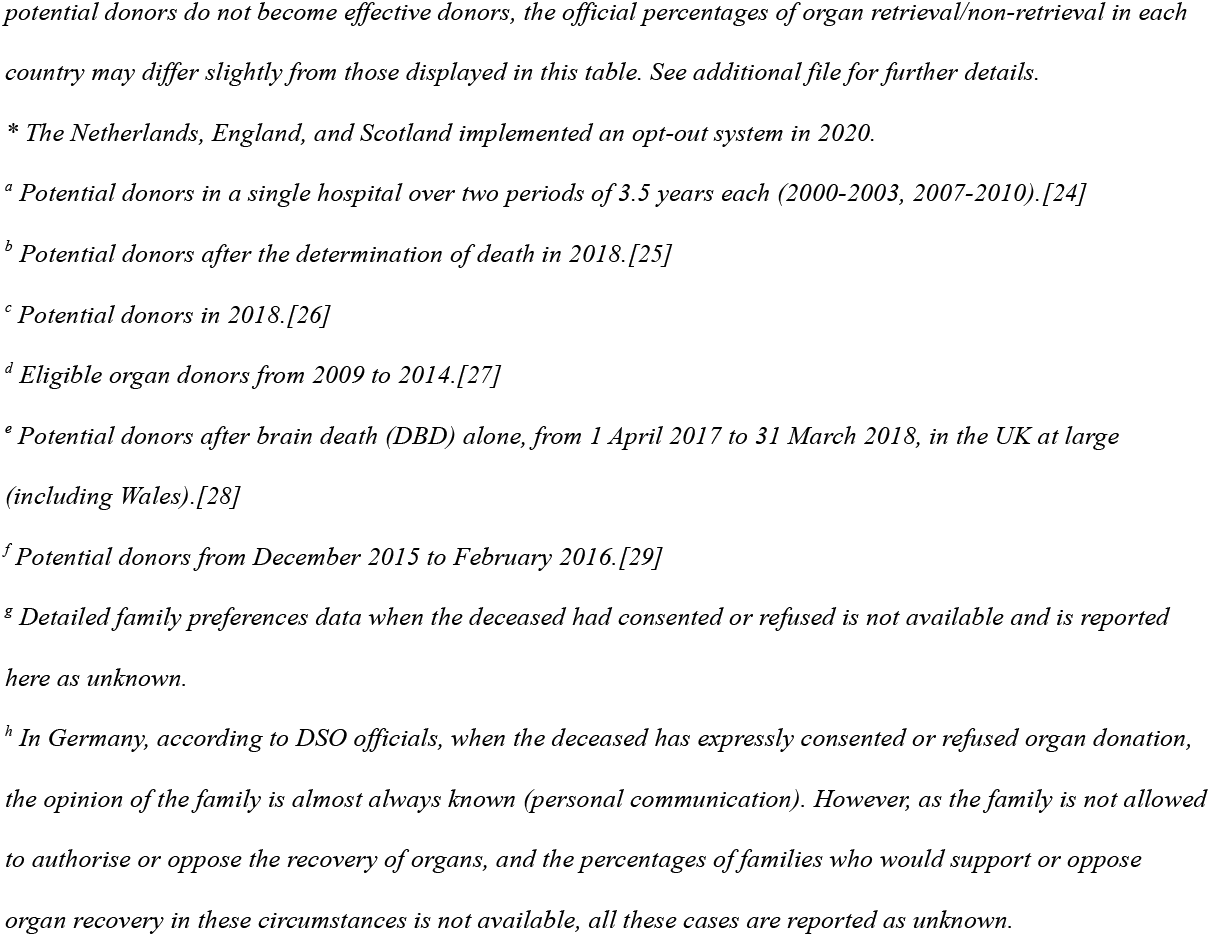
Actual frequency of each scenario among potential organ donor cases when both the deceased’s decision and the family’s preferences are considered in Denmark, Germany, the Netherlands (NL), Sweden, the United Kingdom at large (UK) and Wales in particular.

#### Additional supporting evidence

We found partial statistics from 15 countries regarding the situation where policy defaults apply, that is, when both the deceased and the family preferences are unknown. In particular, we found nationwide statistics from official sources in Belgium, Chile, Colombia, Ireland, Spain, and Switzerland. We also found retrospective studies, mostly from a single hospital and varying periods of time, in Australia, Brazil, Finland, France, Hong Kong, South Africa, South Korea, Spain, Turkey, and the United States. In addition, we obtained informal comments and assessments through personal communication with officials from Belgium, Colombia, Denmark, Finland, South Korea, and Spain. More detailed information about the data, sources, and methodology is available in the Supplementary file.

Results suggest that the differential impact of opt-in and opt-out policies is limited to a range of 0% to 2% of all retrieval opportunities in seven countries (Australia, Belgium, Chile, Colombia, Finland, South Korea, and Spain), to a range of 3% to 5% in five countries (France, Hong Kong, Switzerland, Turkey, and the United States), and to more than 5% in two countries (Brazil and South Africa). These results coming from a wide variety of countries are consistent with those detailed in Table 4.

### Estimation of potential retrieval rates under different policies in seven countries

To better assess the relative impact of family’s intervention in each consent system, we estimated the potential for organ retrieval in four distinct scenarios (Fig. 1). On the one hand (left), we considered opt-in and opt-out policies based on the deceased’s wishes alone, without any family intervention. On the other hand (right), we considered opt-in and opt-out policies based on both the deceased’s and the family’s wishes. In other words, for each reviewed country, we estimated the potential for organ retrieval if the policy in place in that country was: (α) opt-in and deceased’s wishes alone; (β) opt-out and deceased’s wishes alone; (γ) opt-in with family intervention; and (δ) opt-out with family intervention (see the supplementary file for more information about the data and methodology).

**Fig 1:**
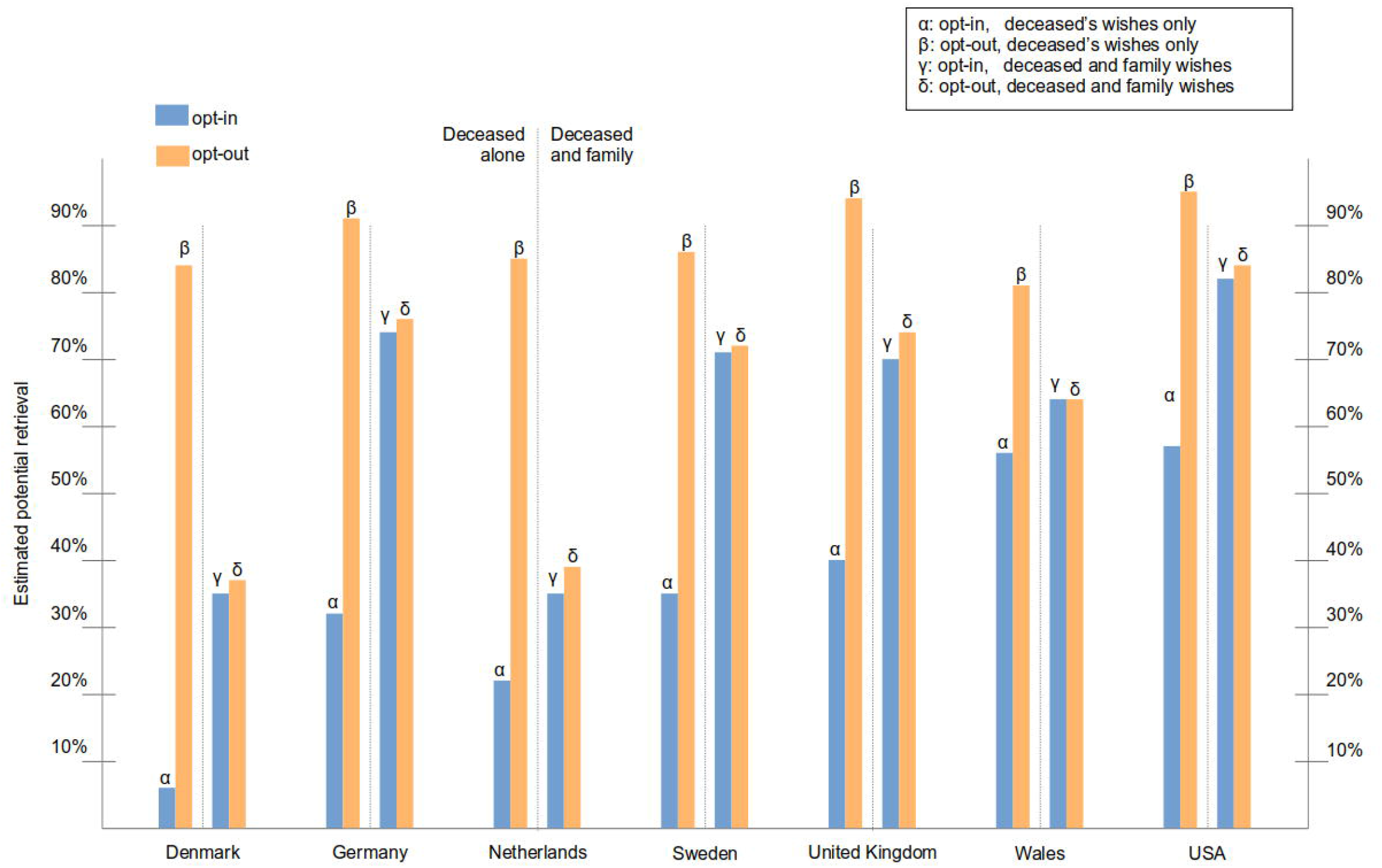
Estimated potential retrieval rates in several countries in four situations depending on the model of consent and the role of the family. For each country, four possible situations are considered, from left to right: (α) opt-in and deceased’s preferences only; (β) opt-out and deceased’s preferences only; (γ) opt-in and both deceased’s and family preferences; (δ) opt-out and both deceased’s and family preferences. Data for this figure results from adding the percentages of the scenarios shown in Table 4 (Denmark, Germany, the Netherlands, Sweden, the United Kingdom, and Wales). In addition, we extracted data from the US National Survey of Organ Donation Attitudes and Practices under the assumption that these figures may somehow reflect actual practice (which is not necessarily the case) to explore how the potential for organ retrieval compares under the four aforementioned situations^38^. For each country, the situation that is actually in place in the country is signalled by an arrow.

The estimated potential retrieval rates in these four scenarios suggest that individual consent policies only make a significant difference when family preferences are disregarded. In this case, moving from opt-in to opt-out may dramatically increase the number of potential donors from which organs can be retrieved (left bars). However, when families are allowed to intervene and their own preferences are taken into consideration, then the potential retrieval outcomes under opt-out are just a little higher than under opt-in (right bars).

## Discussion

Our results shows that the retrieval outcomes under opt-in and opt-out policies are identical in all situations but one, which is when organ donation preferences have not been expressed and, therefore, defaults apply. It is the frequency of this particular situation that determines the impact of consent policies on donation rates, because it determines how often policy defaults apply.

If only the preferences of the deceased person were considered, opt-out would allow the recovery of organs from all individuals who failed to express any preference, whereas opt-in would prevent it. Depending on how often this situation would happen in a given country, moving to opt-in to opt-out could dramatically increase the rates of organ recovery. From an analytical perspective, it would be a 33% increase. In the real world, it is difficult to say, because there is hardly any country in the world where only the preferences of the deceased person are considered[2,17,21,30–32]. Indeed, most opt-in and opt-out countries worldwide allow the family, either *de jure* or *de facto*, to make a decision when the deceased had not, and even to overrule the deceased’s consent to donate (cf. Supplementary file)[8,21,32].

If both the preferences of the deceased and those of the family are considered, then opt-out allows the recovery of organs when neither the deceased’s nor the family’s preferences are known to the medical team. This is obviously a less frequent situation. According to our analysis, if all retrieval opportunities were evenly distributed among all situations, moving to opt-out would increase organ retrieval rates by 11%. In the real world, the impact of the policy is limited to 5% or less. In other words, if any of the reviewed opt-in countries decided to adopt an opt-out policy, organ retrieval would potentially increase by 0% to 5%. Conversely, if any of the reviewed opt-out countries decided to adopt an opt-in policy, organ retrieval would potentially decrease by 0% to 5%.

Our study also shows that the intervention of the family improves organ retrieval under opt-in but hinders it under opt-out (Fig. 1). Though this may seem counter-intuitive, a plausible explanation for this phenomenon is the following. The intervention of the family increases the proportion of likely organ donors under opt-in policies (Fig. 1, blue bars) in all examined countries, as family authorisations in absence of the deceased’ consent outnumber family oppositions when the deceased had consented. In other words, as a majority of deceased individuals fail to express their preferences before death, a majority of organs could not be retrieved in opt-in countries but for the next-of-kin’s authorisation. Meanwhile, family intervention decreases the proportion of likely organ donors under opt-out policies (Fig. 1, orange bars). Indeed, when the deceased consented or their preferences are unknown, family oppositions prevent the retrieval of organs that would otherwise be retrieved. In other words, the organs of all those who remained silent could be retrieved in opt-out countries if it was not because of opposition from families.

The power of our approach stems from the combination of analytical methods with real-world statistics from multiple and diverse countries, allowing us to measure the frequency of that particular situation where opt-in and opt-out policies actually differ in their application. In other words, our study is the first to examine the impact of opt-in and opt-out by focusing on what makes these policies different from each other. To our knowledge, this specific information has never been actively sought nor specifically published before in the scientific literature, and it is seldom reported in official statistics even in countries, such as Spain, with advanced organ donation and transplantation programmes. This makes the data we obtained the best empirical evidence available to date.

The main caveat of our study is the heterogeneity of sources, sample sizes and time periods for the data collected, especially for the additional supporting evidence. However, we were reassured by the fact that, despite this heterogeneity, results converge: across different countries from the five continents, in different hospitals and different time frames, the only situation where consent policy defaults actually apply is rather uncommon.

This study only considers the direct effects, but not the indirect effects of opt-out policies on organ retrieval rates. For instance, presumed consent may enable the initiation of organ preservation measures when the preferences of the deceased and those of the family are absent. This could explain the higher prevalence of uncontrolled donation after circulatory arrest protocols –which require expeditious organ preservation measures– in opt-out countries as compared to opt-in countries[33,34]. Opt-out legislation may also have a psychological influence, either positive or negative, on prospective candidates for organ retrieval and their families[35,36]. It may alter the way in which health professionals talk to families, as well as conversations between family members[5]. In Wales, a statistically significant increase in the proportion of relatives authorising organ retrieval, 33 months after the introduction of the opt-out system, could be explained by such indirect factors, which include familiarity with the legislation, training, increased societal concern about organ scarcity, and growing confidence of families in healthcare professionals[37]. However, the introduction of opt-out legislation has also had the contrary effect, with people rushing to register themselves as non-donors in Brazil and family oppositions skyrocketing in Chile[38,39]. Whether and to what extent consent policies have an influence on family authorisation or refusal rates requires further investigation. The governance quality of these policies also requires further investigation[40].

Our results may warn contemporary organ retrieval policy makers that, by emphasizing the need to introduce presumed consent, they might be overestimating the influence of policy defaults, and underestimating the power granted to families in expressing their preferences and making decisions about organ donation. Improving infrastructures, coordination and training, communication to the public, and modifiable factors influencing family authorisation might prove more effective for increasing organ retrieval rates than moving from opt-in to opt-out.

## Data Availability

All data referred to in the manuscript were openly available prior to the initiation of the study. Data sources used for this study are cited in the manuscript or in the supplementary file.

## Acknowledgments

The authors thank the following persons for their assistance in gathering or interpreting specific data: Lone Bøgh (Dansk Center for Organdonation, Denmark), Beatriz Domínguez Gil (ONT, Spain), Magdalena Flatscher-Thöni (Tyrolean Private University, Austria), Solveig Lena Hansen (University of Bremen, Germany), Anna-Maria Koivusalo (Helsinki University Hospital, Finland), Axel Rahmel (DSO, Germany), Jeantine Reiger-Van de Wijdeven (NTS, Netherlands), Gabriele Werner-Felmayer (Medical University Innsbruck, Austria), Sabine Wöhlke (University of Göttingen, Germany), Won-Hyun Cho (Korea Organ Donation Agency, South Korea), Zeynep Ugur (Social Sciences University of Ankara, Turkey), Kristof Van Assche (University of Antwerp, Belgium), Britzer Paul Vincent (University of Bedfordshire, UK), and Stela Zivcic-Cosic (University of Rijeka, Croatia), as well as the staff of INCUCAI (Argentina), Instituto Nacional de Salud (Colombia), Agence de la Biomédecine (France), Nederlandse Transplantatie Stichting (the Netherlands), and NHS Blood and Transplant (UK) for answering our questions and checking or discussing our results. The authors also thank the following persons for their comments: Anne Dalle Ave (University Hospital of Lausanne, Switzerland), Dale Gardiner (Nottingham University Hospitals, UK), Walter Glannon (University of Calgary, Canada), Alicia Pérez Blanco (ONT, Spain), Gurch Randhawa (University of Bedfordshire, UK), and Stuart Youngner (Case Western Reserve University, USA).

## Ethics statement

All datasets used in this study were publicly available prior to the initiation of the study. No IRB or ethics committee approval was required.

## Data sharing statement

All data used in this study were publicly available prior to the initiation of this study. The sources and analysis of the data used are openly available as supplementary material at medRxiv (doi: https://doi.org/10.1101/2021.08.27.21262033). Requests for further information can be sent to the corresponding author.

## Contributorship statement

The study concept was conceived by AMP, DRA, and JD. The search and analysis of data was conducted by AMP. The manuscript was drafted by AMP and DRA, and critically revised by JD. All authors read and approved the final version of the manuscript.

