## Supplementary file for "Opt-out policies capacity to increase organ donors is limited"

### Supplementary information

Molina-Pérez, Alberto[1,2,3]*; Rodríguez-Arias, David[2,3]; Delgado, Janet[2,3]

1 Institute for Advanced Social Studies, Spanish National Research Council, Spain.

2 Department of Philosophy 1, Faculty of Philosophy, University of Granada, Spain.

3 ESOT-ELPAT Public Issues Working Group.

#### 1. Differential impact of opt-in, opt-out policies when only the deceased´s preferences are considered

A 2012 review of 27 nations where organ donation registries exist showed that only 141 million people out of 719 million (20%) were registered worldwide [1]. In Europe, among 16 nations with registries, only 27 million people out of 272 million (10%) had registered a decision in favour or against donation [1]. As regards organ donor cards, a 2006 survey of 25 EU member states reflected a similar average: only 12% of European citizens were card holders [2]. A further survey of 27 EU member states in 2009 showed that 40% of Europeans had raised the issue of organ donation and transplantation with their family, compared to 59% who had never broached the subject [3].

These figures may have improved over time with variable intensity across countries and procedures. For instance, accrued registrations slightly improved in the Netherlands from 40% in 2010 [1] to 49% in January 2020 [4]. In Germany, the total number of donor-card holders rose from 9% in 2006 [2] to 36% in 2019 [5], and family discussions from 44% in 2009 [3] to 47% in 2019 [5]. Overall, 56% of German citizens had made a decision in 2019, but only 47% had documented and/or communicated it to someone [5]. In the UK, accrued registrations went from 37% in 2009 [3] to 38% in 2019 [6]. In Belgium, the total number of registered citizens increased from 300,000 in 2010 [3] to 550,000 in 2018 [7], which still represents less than 5% of the population. In Italy, cumulative registrations doubled from 1.2 million in 2010 [1] to 2.5 million in 2019 [8], which is nearly 4% of the population. In France, the refusal register is by law the main procedure to express a decision, yet only 300,000 people –less than 0.5% of the total population– were listed in the register by 2017 [9,10].

#### 2. Classification of the role of the family in opt-in, opt-out countries

**Table 1. Classification of the role of the family in opt-in, opt-out countries according to Delgado and colleagues (2019) fourfold level of involvement[11].** Depending on each country's policy, the family may have either no role at all in the decision-making process (L0), act as a mere witness to the deceased's wishes, without any capacity to make a decision (L1), act as a surrogate decision-maker when the deceased had not expressed any preference (L2), or have full decisional capacity, including the capacity to overrule the deceased's explicit wishes (L3). Delgado and colleagues differentiate the family's involvement according to the law and in clinical practice. The other two sources are not specific enough about this difference. A previous and slightly different version of this table has been published elsewhere[12].

<table>
<tr><th></th><th></th><th colspan="4">Role of the family</th><th></th></tr>
<tr><th>Consent policy</th><th></th><th>L0<br>No role</th><th>L1<br>Witness</th><th>L2<br>Surrogate</th><th>L3<br>Full decisional capacity</th><th>Source</th></tr>
<tr><td rowspan="4">opt-in</td><td>law</td><td></td><td></td><td>CA, DE, NL[a], UK[b], USA</td><td>IN</td><td rowspan="2">[11]</td></tr>
<tr><td>practice</td><td></td><td></td><td>DE</td><td>CA, IN, NL[a], UK[b], USA</td></tr>
<tr><td></td><td></td><td></td><td>NL, RO, UK, USA</td><td>AU, BR, CA, CH, CU, DE, DK, EE, IE, IL, IN, IS, JP, KR, KW, LT, MT, MX, MY, NZ, PH, SA, TH, VE, ZA</td><td>[13]</td></tr>
<tr><td></td><td colspan="2">CA</td><td></td><td>AU, CH, DE, DK, IE, IL, NL, NZ, UK, USA</td><td>[14]</td></tr>
<tr><td rowspan="4">opt-out</td><td>law</td><td>AR, AT, PT[c], UY</td><td>BE[d], CL, ES, FR, SG</td><td>SE</td><td>JP</td><td rowspan="2">[11]</td></tr>
<tr><td>practice</td><td></td><td></td><td>BE, SE, SG</td><td>AT, CL, ES, JP, NO, PT</td></tr>
<tr><td></td><td></td><td></td><td>BE, FI, SG, SE</td><td>AM, AT, BY, CL, CO, CR, CZ, EC, ES, FR, HR, IT, LU, NO, PL, PY, RU, SI, SK, TN, TR</td><td>[13]</td></tr>
<tr><td></td><td colspan="2">AT, CZ, LU</td><td>GR, PT, SK</td><td>BE, ES, FI, FR, HR, HU, IT, NO, RO, SE, SI</td><td>[14]</td></tr>
</table>

AM: Armenia; AR: Argentina; AT: Austria; AU: Australia; BE: Belgium; BR: Brazil; BY: Belarus; CA: Canada; CH: Switzerland; CL: Chile; CO: Colombia; CR: Costa Rica; CU: Cuba; CZ: Czech Republic; DE: Germany; DK: Denmark; EC: Ecuador; EE: Estonia; ES: Spain; FI: Finland, FR: France; HR: Croatia; HU: Hungary; IE: Ireland; IL: Israel; IN: India; IS: Iceland; IT: Italy; JP: Japan; KR: South Korea; KW: Kuwait; LT: Lithuania; LU: Luxemburg; MT: Malta; MX: Mexico; MY: Malaysia; NL: The Netherlands; NO: Norway; NZ: New Zealand; PH: Philippines; PL: Poland; PT: Portugal; PY: Paraguay; RO: Romania; RU: Russia; SA: Saudi Arabia; SE: Sweden; SG: Singapore; SI: Slovenia; SK: Slovakia; TH: Thailand; TN: Tunisia; TR: Turkey; UK: United Kingdom (Wales excepted); USA: United States of America; UY: Uruguay; VE: Venezuela; ZA: South Africa.

a. The Netherlands has enacted an opt-out policy by July 2020.

b. England and Scotland have implemented opt-out policies by May 2020, and Wales by December 2015.

c. The role of the family in Portugal according to the law was not initially included in Delgado et al. (2019).

d. The role of the family in Belgium according to the law was wrongly classified as *L2: surrogate* in Delgado et al. (2019).

#### 3. Family oppositions

Table 2. ***Family oppositions to organ retrieval in 2016***

| Europe | | | America | | | Other countries | | |
|---|---|---|---|---|---|---|---|---|
| Country | Oppositions (N° of interviews) | Rate (%) | Country | Oppositions (N° of interviews) | Rate (%) | Country | Oppositions (N° of interviews) | Rate (%) |
| Austria | 80 (294) | 27 | Argentina | 434 (1,177) | 37 | Australia | 428 (1,074) | 40 |
| Belgium | 51 (402) | 13 | Brazil | 2,561 (5,921) | 43 | Israel | 52 (137) | 38 |
| Croatia | 20 (186) | 11 | Chile | 139 (272) | 51 | Malaysia | 168 (227) | 74 |
| Germany | 217 (1,399)[a] | 16 | Colombia | 286 (714) | 40 | Saudi Arabia | 230 (333) | 69 |
| Hungary | 16 (242) | 7 | Cuba | 12 (161) | 8 | South Korea | 884 (1393)[b] | 63 |
| Ireland | 36 (72) | 50 | Dominican Republic | 40 (72) | 56 | Turkey | 1,425 (1,988) | 72 |
| Italy | 807 (2,488) | 32 | Uruguay | 2 (96) | 2 | TOTAL | 3,187 (5,152) | 62 |
| Lithuania | 26 (103) | 25 | TOTAL | 3,474 (8,413) | 41 | | | |
| Poland | 71 (677) | 11 | | | | | | |
| Romania | 57 (352) | 16 | | | | | | |
| Slovakia | 11 (112) | 10 | | | | | | |
| Slovenia | 13 (55) | 24 | | | | | | |
| Spain | 372 (2,391) | 16 | | | | | | |
| UK | 1,172 (3,145) | 37 | | | | | | |
| TOTAL | 2,949 (11,918) | 25 | | | | | | |

*Data shows the raw numbers of family oppositions versus total interviews (requests) conducted Only countries where more than 50 interviews were conducted are reported here. Source: EDQM,* Newsletter Transplant 2017.

[a] *Source for Germany: [15].*

[b] *Source for South Korea: [16].*

#### Differential impact of opt-in, opt-out policies when both the deceased's and family preferences are considered

To determine the frequency of each of the nine possible scenarios when both the deceased's and family preferences are considered (see Table 3 of the main article), it is necessary to get very detailed data. We conducted a non-systematic search on Pubmed, Google Scholar, and ResearchGate using the following keywords in three languages (English, Spanish, French): potential donor(s), potential organ donor(s), organ donor audit, potential organ donation, organ donation activity, organ donation referral, organ donation statistics, organ transplantation statistics, and the MeSH terms "Tissue and Organ Procurement/statistics and numerical data" and "Organ Transplantation/statistics and numerical data". We also searched directly into the websites of several national transplant organisations, when available.

We specifically but not exclusively sought data from the following list of 55 countries including most of the statistical sub-regions as defined by the United Nations geoscheme: Algeria, Argentina, Australia, Austria, Belarus, Belgium, Brazil, Canada, Chile, Colombia, Cuba, Croatia, Czech Republic, Denmark, Ecuador, Egypt, Estonia, Finland, France, Germany, Greece, Hong Kong, Hungary, Iceland, India, Iran, Ireland, Israel, Italy, Japan, Kazakhstan, Kenya, Lithuania, Malaysia, Mexico, the Netherlands, New Zealand, Nigeria, the Philippines, Portugal, Romania, Saudi Arabia, Singapore, Slovenia, South Africa, South Korea, Spain, Sweden, Switzerland, Thailand, Turkey, the United Kingdom (in general) and Wales (in particular), the United States, and Uruguay. These 54 countries account for more than two thirds of the 70+ countries having reported some activity in deceased organ donation in recent years to the Global Observatory on Donation and Transplantation database (coordinated by the World Health Organization and the Spanish Transplant Organization) or to the International Registry in Organ Donation and Transplantation (IRODaT.org).

We contacted scholars and officials from the following countries to request data or clarifications:

Table 3: Data requests by email

| Country | Contact person/institution | Date of data request | Answer received |
|---|---|---|---|
| Argentina | INCUCAI (Instituto Nacional Central Único Coordinador de Ablación e Implante) | 18/12/2020 | Yes |
| Australia | DonateLife, Organ and Tissue Authority | 28/01/2021 | No |
| Austria | Magdalena Flatscher-Thöni, Tyrolean Private University; Gabriele Werner-Felmayer, Medical University Innsbruck | 14/01/2021 | Yes |
| Belgium | Kristof Van Assche, University of Antwerp | 14/01/2021 | Yes |
| Colombia | Coordinación Red Nacional de Donación y Trasplantes, INS (Instituto Nacional de Salud) | 02/01/2021 | Yes |
| Denmark | Anja Marie Bornø Jensen, University of Copenhagen | 08/01/2121 | Yes |
| Denmark | Lone Bøgh, Danish Center for Organ Donation | 28/01/2021 | Yes |
| Finland | Findata – Health and Social Data Permit Authority | 04/01/2021 | Yes |
| Finland | Ana Maria Koivusalo, Helsinki University Hospital | 04/01/2021 | Yes |
| France | Agence de la Biomédecine | 04/01/2021 | Yes |
| Germany | Solveig Lena Hansen, University of Bremen | 01/08/2020 | Yes |
| Germany | Axel Rahmel, DSO (Deutsche Stiftung Organtransplantation) | 13/08/2020 | Yes |
| Hungary | Organ Coordination Office | 04/01/2021 | No |
| India | Vasanthi Ramesh, NOTTO (National Organ & Tissue Transplant Organisation) | 04/01/2021 | No |
| Israel | ADI, National Transplant Center | 14/01/2021 | No |
| Israel | Hagai Boas, The Van Leer Jerusalem Institute | 29/01/2021 | Yes |
| Kenya | Ministry of Health | 15/01/2021 | No |
| Lithuania | National Transplant Bureau under the Ministry of Health | 23/01/2021 | No |
| Malaysia | Diana Mohd Shah, National Transplant Resource Center, Kuala Lumpur Hospital | 19/01/2021 | Yes |
| The Netherlands | Jeantine Reiger-Van de Wijdeven, NTS (Nederlandse Transplantatie Stichting) | 26/02/2020 | Yes |
| Philippines | National Kidney and Transplant Institute | 15/01/2021 | No |
| Poland | Poltransplant | 28/01/2021 | No |
| South Korea | Won-Hyun Cho, Korean Organ Donation Agency | 22/01/2021 | Yes |
| South Korea | Claire Junga Kim, Ewha Womans University | 22/01/2021 | No |

| | | | |
|---|---|---|---|
| Spain | Beatriz Domínguez Gil, ONT (Organización Nacional de Trasplantes) | 17/08/2020 | Yes |
| Thailand | Organ Donation Center, Thai Red Cross Society | 24/01/2021 | No |
| United Kingdom | NHS (National Health Service) | 27/02/2020 | Yes |
| Uruguay | INDT (Instituto Nacional de Donación y Trasplante de Células, Tejidos y Órganos) | 02/01/2021 | No |

**In the following, scenario *Cc* refers to the particular situation where both the deceased's preferences and the family's preferences are unknown to the medical team.** Family preferences can remain unknown under several circumstances, including: (a) the deceased had no living relatives or close friends to be asked; (b) relatives could not be contacted in time; (c) relatives were too distressed to be asked or to make a decision regarding donation; (d) relatives held conflicting views and could not reach a common decision.

Eventually, we found partial data regarding scenario *Cc* (when both the deceased's preferences and the family's preferences are unknown) in Australia, Brazil, Belgium, Chile, Colombia, Denmark, Finland, France, Hong Kong, Ireland, South Africa, South Korea, Spain, Switzerland, Turkey, and the USA. We also found more comprehensive data that allowed us to determine the frequency of each scenario in Denmark, Germany, the Netherlands, Sweden, the UK (in general) and Wales (in particular). All datasets used in this study were publicly available prior to the initiation of the study. All data sources are cited below.

**Argentina** (opt-out) implemented in August 2018 a law to limit family's capacity to oppose organ. Official data shows that, between the first and second semester of 2018, family opposition dropped from 43% to 22% and organ donors increased from 279 to 422 [17]. Meanwhile, organ donors increased from an average of 13,1 organ donors per million people (ppm) in the 5-year period (2013-2017) before the law was implemented, to 15,75 ppm in 2018 and a record 19,65 ppm in 2019 [18]. Despite contacting with officials at INCUCAI (the national agency of organ donation and transplantation), we were unable to determine the frequency of scenario *Cc*.

In **Australia** (opt-in), among a cohort of 116 patients admitted at the ICU for "potential organ donation" at The Alfred Hospital, Melbourne, from 2007 to 2016, organ donation could not be discussed with the family in only one case [19]. In all other cases, the family could make a decision. This means that scenario Cc occurred in 1% of cases in that cohort during a 10-year period.

In **Belgium** (opt-out), there was 712 potential organ donors in 2019 and the reasons for non-procurement were: 38,3% medical contraindications, 12,6% family refusal, 1,3% registered refusal, 0,4% objection by the coroner, and 4,4% because of "other/unknown" reasons [20]. If we exclude all cases of medical contraindications, there were 439 potential donors in 2019, out of which 31 (7%) were not retrieved because of "other/unknown" reasons. However, these cases may or may not correspond to scenario Cc. Therefore, cases of scenario Cc may be as low as 0% or as high as 4% or 7% of cases, depending on the definition of potential donor. When searching further information, we were told by a Belgian representative in the European Committee on Organ Transplantation that "The situation where the potential donor had failed to make a decision and the preferences of the family regarding donation are also unknown, is extremely rare. If the family cannot be reached after trying for a long time, the opt out will be performed. For Belgium I know maybe of 2 cases in my whole career where this happened. If the potential donor had failed to make a decision and the family can be reached, we only contact the family members in the first degree. If they are disagreeing among themselves, we leave them some time and then re-contact them. If there is still disagreement, we will not proceed with the organ removal" (Personal communication). Therefore, although the number and ratio of cases of scenario Cc is not known, available data indicates that these cases are likely very rare and that, in any case, they were below 4-7% in 2019.

In **Brazil** (opt-in), a retrospective study in a single hospital between January 2008 and December 2010 found 1346 deaths, with 41 notifications of brain death, 21 notifications of cardiac death (out of 1305 cardiac death), and a total of 1284 non-notifications, out of which 49 cases were not notified because of "patients without identification" and 11 cases because of "missed family" [21]. This means that scenario Cc would represent 49+11 out of 1346 cases, that is 4.3%. However, it is not clear to us that all deaths reported here are cases of potential donors. A different study of 47 records of family refusal from an hospital in the state of Parana reports 5 cases of family disagreement [22]. However, the number of potential donors in this study is not mentioned. Another retrospective study in an area of Brazil's Northest between Nov. 2015 and Jan. 2017 found 150 brain dead potential donors and 74 cases of family refusal, out of which 17 because of the deceased's refusal in life and 15 because of familiar disagreement [23]. If we consider these cases of familiar disagreement as instances of unknown wishes, instead of instances of family refusal, then scenario Cc would have occurred in 10% of cases in that area.

In **Chile** (opt-in in 2008), the 2008 annual report of the now extinct Transplant Corporation (Corporación del Trasplante) indicates that, out of 303 potential donors, the family could not be contacted in two occasions and the deceased had no family in one occasion [24]. This means that scenario Cc occurred in 1% of total cases in 2008.

In **Colombia** (opt-out), presumed consent for organ donation has been implemented in 1988 and the family was allowed to authorised or oppose donation. This law was amended in 2016 so that the family is no longer allowed to intervene (to authorise or to oppose) since 2017. However, the number of family authorisations, oppositions, and cases of presumed consent were recorded up to (and including) 2017 (see Table below). Since 2018, the occurrences of family acceptance and refusal are no longer recorded in official statistics because the 2016 law does not allow the family to intervene in the decision (Coordinación Red Nacional de Donación y Trasplantes: personal communication on January 19$^{th}$, 2021). The table below shows that during the four years before the new law was enacted, presumed consent was anecdotal (0% to 1%) and rose to 4% in 2017 (data source: [25]).

Table 4: Organ donation statistics in Colombia up to 2017

| Year | 2013 | 2014 | 2015 | 2016 | 2017 | Total |
|---|---|---|---|---|---|---|
| Eligible donors | 621 | 662 | 823 | 715 | 755 | 3576 |
| Family opposition | 229 | 254 | 325 | 286 | 203 | 1297 |
| Family authorisation | 392 | 408 | 493 | 428 | 521 | 2242 |
| Presumed consent | 1 | 1 | 5 | 1 | 31 | 39 |
| Total donors | 393 | 409 | 498 | 429 | 474 | 2203 |

Source: Colombia's Instituto Nacional de Salud [25].

In **Finland** (opt-out), presumed consent has been implemented in 2010 and the family is not legally allowed to oppose organ donation. However, in practice, organs are never procured against the wishes of the family (National transplant coordinator, Hospital University of Helsinki: personal communication). Data from a single hospital study between 2005 and 2015 identified 83 potential organ donors with no case corresponding to scenario *Cc* [26]. Finland's national transplant coordinator confirmed to us that it is extremely rare that the family cannot be contacted and this situation may have happened only once in several decades (personal communication). This means that the frequency of scenario Cc is nearly 0%.

In **France** (opt-out), a law to limit the family's capacity to oppose organ donation has been implemented on January 1st, 2017. Since that date, the family cannot legally oppose organ donation but only act as a witness of the deceased's wishes (when the deceased did not register their refusal). Official data shows that, in the 3-year period before (2014-2016) and after (2017-2019) the law was implemented, oppositions (either by the deceased or the family) dropped by 2,2% from 32,5% to 30,3%, and organ donors increased by 2,2% from 48% to 50.2% of all brain dead patients [27]. According to a Parliament's internal report in December 2017, relatives still opposed organ donation *de facto*, although this was not allowed *de jure*, either by claiming (wrongfully) that the deceased was unwilling to donate while alive, or by expressing fierce or even violent [sic] objections to the medical team [10]. In the latter case, this is officially reported as "because of the context, organ procurement was not possible" ("en raison du contexte, le prélèvement n'a pas été possible") which is a new category created in 2017. Official records for 2019 report that, out of 3,471 brain dead individuals, there were 1,729 procurements, 683 non-procurements for reasons unrelated to refusal/opposition (mostly medical contraindications), and 1,059 non-procurements because of oppositions classified as "opposition from the legal representative" (N=50), "because of the context,…" (N=491), and "deceased's refusal" (N=518) (source: Agence de la biomédecine, https://rams.agence-biomedecine.fr/le-prelevement-dorganes-en-vue-de-greffe, "Prélèvement sur donneur décédé en état de mort encéphalique",

Tableau P2 & P7). There is no direct indication of situations that would correspond to scenario *Cc* and we were unable to determine or estimate the frequency of that scenario from official data. A representative of the Agence de la Biomédecine confirmed to us that there is no official data available and that only transplant coordinators may know from their own personal experience the frequency of scenario Cc (personal communication). Nonetheless, we managed to find two studies from a single harvesting centre that mention family disagreement: 3 cases out of 227 potential organ donors between Jan. 2010 and Dec. 2011 and 22 out of 426 between Jan. 2012 and Dec. 2015, respectively 1% and 5%, although they do not mention if a decision was made eventually by the family [28,29]. These two studies were conducted by the same authors, in the same centre, with the same methodology, during consecutive periods. Therefore, by combining their results, we have 3+22=25 cases out of 227+426=653 potential donors: 3.8%.

In **Hong Kong** (opt-in), a study of all family members of potential donors after cardiac death approached by the local eye bank staff members from January 2008 to December 2014 identified 1,740 cases, out of which 1,099 refused corneal donation and 79 of them because of “lack of consensus in the family” [30]. Since only families approached are included, this means that scenario Cc occurred in at least 4.5% of cases. Another retrospective study conducted in 2014 at Queen Elizabeth Hospital, the largest regional acute hospital in HK, reported 21 eligible donors after brain death and families were approached in all cases, which means that scenario Cc did not happen [31].

In **Iceland** (opt-out), a study of all brain dead patients in two hospitals from Jan. 2003 to Dec. 2016 mentions 125 potential donors and 99 eligible donors, with 64 cases of family consent, 21 cases of family refusal, and 15 cases of permission not requested, among which 11 cases where the reason for not requesting permission for donation could not be determined [32]. If all these cases were instances of scenario Cc (which we actually don’t know), this scenario would be limited to 11% or less of all eligible donors.

In **India** (opt-in), data from a single hospital reported 61 potential donors after brainstem death, out of which “based on their previous interaction with the patients’ family members, the treating team did not feel it appropriate to counsel four patients’ family members for deceased organ donation” [33]. However, it is not clear if the medical team did not approach the family because of their emotional distress or because they were likely to refuse.

In **Iran** (opt-in), a study of the causes of family refusal in 2009-2010 (73 out of 125 brain dead patients) mentions 3 cases as “other” [34]. If these were all cases of scenario Cc (which we don’t know), this scenario would be limited to 4% or less.

In the **Republic of Ireland** (opt-in), data from a potential donor audit shows that, between September 2007 and August 2008, among 138 potential donors, when the deceased had not expressed any preference, the family preferences remained unknown in four cases, namely, because the next-of-kin was not traceable in time, the family was too distressed, there was a language barrier, and one unspecified reason [35,36]. This means that scenario Cc occurred in 3% of all potential donor cases during that period.

In **South Africa** (opt-in), data from the Groote Schuur Hospital, Cape Town, over a 10-year period (2007-2016), indicates that, among 861 patients referred for donation, the identity of the patient was unknown or their family could not be contacted successfully in 72 cases [37]. This means that scenario Cc occurred in 8.4% of all cases in that hospital during that period.

In **South Korea** (opt-in), data from a retrospective survey (2014-2019) shows that, among 9,587 potential brain death cases suitable for organ donation, 536 did not proceed because of disagreements between family members, which represents less than 6% of the total [38]. This is consistent with a second study (2013-2018) indicating that out of 6,987 medically available potential brain deaths, 441 (6.3%) were initially consented by next of kin but refused by other family members [16]. However, the first survey cited provides no detailed information (it is just a conference abstract) and the second survey cited shows contradictions: Table 2 indicates 105 cases of initial consent by next-of-kin but refusal by other family members (in 2018), while Fig. 1 indicates 100+5 cases mentioning respectively “Refused by medical staff” and “No family member for consent”, in addition to 11 cases of “cancel consent”. A third study using data from the same period (2013-2018) indicates 100 cases mentioning “Refused to meet medical staff” and 3 cases of “donation withdrawal” (in 2018) [39]. In addition, while the second study mentions that, out of 884 cases of donation refusal in 2018, 655 families refused the interview itself when approached [16], the third study mentions 88 cases of donation refusal in 2018 and 100 additional cases of families that “refused to meet medical staff” [39]. In Korea, the authorisation of the family is mandatory for organ donation, and refusal to meet the medical staff may be considered a way to oppose organ donation. With regard to family disagreement, this may or may not be understood as a situation of unknown family wishes because some family members initially accept organ donation and that decision is eventually overruled by other family members, so that the final decision of the family is a refusal. We contacted the author of the second study, who is the president of the Korea Organ Donation Agency, for clarification. Based on all this, we consider that the only clear cases of unknown family wishes are those where no next-of-kin is available to make a decision. This corresponds to 30 cases out of 1,833 medically available potential donors in 2018, that is 1.6% [16].

In **Spain** (opt-out), the data on outcomes from potential donors recorded by the National Transplant Organization (ONT) is not detailed enough to ascertain the relative frequency of all nine scenarios or of scenario *Cc*. A study of 1,844 brain dead patients from 42 ICUs mentions 7 cases of patient’s refusal during lifetime, 244 cases of family opposition, and no causes of loss of donors that can correspond to scenario Cc beside 14 cases classified as “other” [40]. If all these 14 “other” cases were instances of scenario Cc, this scenario would occur in less than 1% of potential donor cases. Another study at a high-volume, tertiary hospital with one of the highest proportions of DCD in the country mentions that, out of 621 donor-eligible individuals from 2008 to 2017, 5 were lost owing to the absence of a decision-making family member [41]. These cases corresponds to the definition of scenario Cc and represent less than 1% of eligible donors. ONT statistics relative to donation after brain death show that, in 2018, there were 1,607 organ donors, 313 cases of family refusals, and 5 cases in which the family could not be contacted [42]. This means that the frequency of scenario Cc is nearly 0%. ONT authorities confirmed to us that scenario Cc is extremely rare (personal communication).

In **Switzerland** (opt-in), data from Swisstransplant shows that, between September 2011 and August 2012, among 216 potential donors, when the deceased failed to express any preference, the next-of-kin could not be contacted in six cases, and the family could not make a decision in one case [43–45]. This means that scenario Cc occurred in 3% of all potential donor cases during that period.

In **Turkey** (opt-out), a study of all adult and paediatric patients diagnosed with brain death between January 2001 and December 2016 in a tertiary reference university hospital with 73 ICU beds shows that, out of 303 patients with brain death, the patients’ relatives could not be interviewed (no reason mentioned) in 5 occasions [46]. If we suppose that these families could not be contacted, the frequency of scenario Cc would be 2%. Data from another single hospital study between 2008 and 2019 mentions that, out of 82 brain dead potential donors, 3 cases of families who refused to communicate for the donation process [47]. This corresponds to 3.7%.

In the **United States of America**, a retrospective analysis of all referrals during a 4-year period (2004-2007) to an Organ Procurement Organization (OPO) in South and Central Texas mentions that 44 families of the 827 potential organ donors referred to that OPO were divided on the decision [48]. No indication is given on whether the deceased had expressed their willingness to donate. This means that scenario Cc occured in 5% or less of all potential donor cases referred to that OPO during that period. In the USA at large, a review of hospital medical records of deaths occurring in the ICU from 1997 through 1999 in 36 OPOs indicates that “in 3 percent of the cases, organs were not donated for other reasons, such as restrictions imposed by the medical examiner, the occurrence of cardiac arrest in the patient, precluding organ recovery, or the lack of a family member who could give consent.” [49]. A study of donation decisions of 420 families in Ohio and Pennsylvania identified 26 cases of family disagreement [50].

Table 5. ***Estimated frequency of scenario Cc in several countries.***

| Consent model | Country | Period | Potential/eligible donors[a] | Cases of scenario Cc (%) |
|---|---|---|---|---|
| Opt-in | Australia | 2007-2016 | 116[c] | 1 (1) |
| | Brazil | 2016 | 150[c] | 15 (10) |
| | Hong Kong | 2008-2014 | 1740 | 79 (5) |
| | Rep. of Ireland | 2007-2008 | 138 | 4 (3) |
| | South Africa | 2007-2016 | 861[c] | 72 (8) |
| | South Korea | 2018 | 1,833 | 30 (2) |
| | Switzerland | 2011-2012 | 216 | 7 (3) |
| | USA | 1997-98 | n/a | n/a (3) |
| | | 2004-07 | 827 | 44 (5) |
| | | n/a (~2005) | 420 | 26 (6) |
| Opt-out | Belgium | 2019 | 712 | n/a (0-4) |
| | Chile[b] | 2008 | 303 | 3 (1) |
| | Colombia | 2013-2017 | 3,576 | 39 (1) |
| | Finland | 2005-2015 | 83 | 0 (0) |
| | France | 2010-2015 | 653[c] | 25 (4) |
| | Spain | 2018 | 1925 | 5 (0) |
| | Turkey | 2001-2016 | 303[c] | 5 (2) |
| | | 2008-2019 | 82[c] | 3 (4) |

[a] The definition of a potential donor varies between countries.

[b] Chile had an opt-in system in 2008 and switched to opt-out in 2010.

[c] Data from a single hospital or a single organ procurement organization.

In addition, we were able to extract the detailed information required for filling the table from six countries: Denmark, Germany, the Netherlands, the United Kingdom, Wales, and Sweden. German colleagues helped us interpreting documentation written in German. Then, we contacted the respective national transplantation organisations by email to confirm our results. We received confirmation from the Nederlandse Transplantatie Stichting (Netherlands) and the NHS Blood and Transplant (UK). The DSO (Germany) answered our email but did not provide confirmation or refutation of our results.

#### Denmark

Source: Thybo KH, Eskesen V. The most important reason for lack of organ donation is family refusal. Dan Med J. 2013;60: A4585. [51]

A retrospective study of all patients who died at Rigshopsitalet between 2000-2003 and 2007-2010 shows that, out of a total of 284 potential donors during these two periods, 25 (12+13) were eventually not converted into actual donors because of contraindications, 23 (12+11) because donation was not considered, and 2 (1+1) because of forensic examination. If we exclude these 49 cases of non-donation that are unrelated to consent, the number of potential donors is 235.

The number of effective donors is 90 (42+48). Among them, 15 (5+10) had registered their willingness to donate, and in the remaining 75 cases, authorisation was given by the next-of-kin (scenario Ca).

The number of non-donors is 167 (98+69). Among them, the deceased had refused donation in 18 (15+3) cases, and the next-of-kin opposed it in 120 (67+53) cases. In addition, there are 6 (4+2) cases signalled as “no next of kin”.

This is consistent with but higher than nationwide data from the Dansk Center for Organ Donation for the period 2014-2018, as mentioned above, with 9 cases where both the wishes of the deceased and those of the family were unknown because relatives could not be found (personal communication). The annual average of potential donors in this period was 260, which means a total of approximately 1300 (personal communication). Therefore, the frequency of scenario Cc in Denmark for that period was less than 1%.

Denmark has an opt-in system where the family can make a decision when the deceased had not, but cannot legally overrule (veto) the deceased’s consent. Therefore, we can consider that all 120 cases of family opposition correspond to scenario *Cb*. When the deceased had made a decision, the preferences of the family are not considered and remain unknown to us. Therefore, the 15 cases of deceased’s consent correspond to scenario *Ac* and the 18 cases of deceased’s refusal correspond to scenario *Bc*.

Table 6

| **N=235 potential donors** | | | **Family preferences** | | |
|---|---|---|---|---|---|
| | | | **(a) favorable** | **(b) unfavorable** | **(c) unknown** |
| | | | *31.9% (N=75)* | *51% (N=120)* | *16.6% (N=39)* |
| **Deceased’s decision** | **(A) consent** | *6.4% (N=15)* | *n/a* | *n/a* | *6.4% (N=15)* |
| | **(B) refusal** | *7.7% (N=18)* | — | *n/a* | *7.7% (N=18)* |
| | **(C) unknown preferences** | *85.5% (N=201)* | *31.9% (N=75)* | *51% (N=120)* | *2.6% (N=6)* |

In addition, a study of all patients who died in Aalborg University Hospital in 2012 shows that 47 patients died in ICU and 32 where identified as potential donors, out of which “3 patients died without having their relatives confronted by ICU staff about organ donation” but no mention is made of the reasons why relatives were not approached, and all other families either authorised (12) or refused (19) organ procurement [52]. Depending on whether or not we consider these three cases as instances of scenario *Cc*, the frequency of scenario *Cc* would be either 0% or 10%.

#### Germany

Source: Deutsche Stiftung Organtransplantation (DSO), Jahresbericht organspende und transplantation in deutschland 2018 [15], pages 52 and following:

Fig. 18, p. 52, shows a total of 2,811 donation-related contacts to the DSO before the determination of total brain failure, out of which 1,395 were excluded at this stage and, among those not excluded, 955 became actual donors and 461 did not.

Fig. 21, p. 54, shows that among those 1,395 exclusions, 498 deceased individuals had previously refused to be organ donors (likely through their donor-card). In addition, there are 30 cases where the conversation with relatives was deemed unreasonable, or there was no consent from the authorized persons, or opposition from the prosecutor.

Fig. 22, p. 56, shows that, after the determination of brain death, the total number of potential donors was 1,416, among which there were eventually:

- 955 effective donors
- 340 non-donors because of the deceased's refusal or family opposition
- 99 non-donors for medical reasons
- 22 non-donors for other reasons (including family interview deemed unreasonable or lack of authorisation from the prosecutor/judge)

Fig. 25, p. 58, shows that the number of decisions made about organ retrieval are divided into 1,037 authorisations and 340 oppositions, totalling 1,377. Decisions are distributed as follows:

- ➔ Authorisations (N=1,037):
  - ☞ Presumed willingness to donate: 45,5% (N=472)
  - ☞ Oral consent: 25.4% (N=263)
  - ☞ Written consent: 17.6% (N=182)
  - ☞ Relatives: 11.6% (N=120)
- ➔ Oppositions (N=340)
  - ☞ Presumed willingness not to donate: 31.2% (N=106)
  - ☞ Oral refusal: 32.1% (N=109)
  - ☞ Written refusal: 4.1% (N=14)
  - ☞ Relatives: 32.6% (N=111)

The number of favourable decisions (1,037) is higher than the number of actual donors (955), which means that there were 1,037 - 955 = 82 cases where organs were not actually removed despite authorisation. Thus, among 99 non-donors for medical reasons, there were 82 cases with a favourable decision and 17 cases with no decision being made either in favour or against. In addition, there were 22 cases of non-donors for other reasons, including family interviews deemed unreasonable and lack of relatives to ask for organ retrieval. This gives us 17 + 22 = 39 cases without a decision. This number corresponds to the difference between the total number of potential donors after brain-death and the number of cases with a decision:
1,416 - 1,377 = 39.

- ➔ No decision (N=39)
  - ☞ 17 excluded for medical contraindications
  - ☞ 22 excluded for other reasons (including family interviews deemed unreasonable, no relatives to consult, no authorisation from the prosecutor)

Germany has an “extended consent” policy, which means that it operates an opt-in policy wherein individuals are encouraged to document their wishes on a donor-card (as there is no register) or inform their relatives. When the deceased had not expressed their wishes in written or oral form, relatives are asked to make a decision based on the presumed wills of the deceased. Only if this presumed will of the deceased cannot be determined do the relatives decide according to their own ideas. This corresponds to the distribution of cases of authorisations and oppositions shown above. In the following, we are going to suppose that the deceased´s expressed decision is sufficient (to proceed or not to proceed with organ removal) and that said decision cannot be overruled by the family. This implies that, whenever the deceased had expressed a decision, the preferences of the family are not taken into account. Hence, when the deceased had expressed a decision, family preferences will be considered as unknown.

Now, we can try to figure out the distribution of cases in our scenarios.

- There are 263 cases of oral consent and 182 cases of written consent, totalling 445 cases of expressed willingness to donate by the deceased. As family preferences are considered unknown, these cases correspond to scenario Ac.
- There are 109 cases of oral refusal and 14 cases of written refusal by the deceased, totalling 123 cases. Again, as family preferences are considered unknown, these cases correspond to scenario Bc.
- There are 472 cases of presumed willingness to donate and 120 cases of relatives’ authorisation based on their own preferences, totalling 592. In both circumstances, the deceased’s wishes are unknown, thus corresponding to scenario Ca.
- There are 106 cases of presumed willingness not to donate and 111 cases of relatives’ oppositions based on their own preferences, totalling 217. In both circumstances, the deceased’s wishes are unknown, thus corresponding to scenario Cb.
- There are 22 cases with no decision being made for non-medical reasons.
- There are 17 cases with no decision being made for medical reasons. In these cases, we are going to suppose that *because of medical contraindications* that excluded these individuals as potential donors, the family interview did not take place.

If we exclude these 17 cases of medical contraindications (before families could be asked), we end up with 1,037 (decisions in favour) + 340 (decisions against) + 22 (without any decision) = 1,399 total cases.

Table 7

| **N=1,399 potential donors** | | | **Family preferences** | | |
|---|---|---|---|---|---|
| | | | **(a) favorable** | **(b) unfavorable** | **(c) unknown** |
| | | | *42.3% (N=592)* | *15.5% (N=217)* | *42.2% (N=590)* |
| **Deceased’s decision** | **(A) consent** | 31.8% (N=445) | *0% (N=0)* | *0% (N=0)* | *31.8% (N=445)* |
| | **(B) refusal** | 8,8% (N=123) | — | *0% (N=0)* | *8.8% (N=123)* |
| | **(C) unknown preferences** | 59.4% (N=831) | *42.3% (N=592)* | *15.5% (N=217)* | *1.6% (N=22)* |

### The Netherlands

Source: Nederlandse Transplantatie Stichting (NTS), Jaarverslag 2018 [53], Table 8.3, page 100

Tabel 8.3: **Uitslag Donorregister en reactie nabestaanden onder herkende pontentiële orgaan-donoren op ic-afdelingen van 87 ziekenhuizen in 2018** (bron: NORD-MSO)

| Uitkomst Donor-register (DR) | Aantal herkende potentiële donoren | % van alle raadplegingen met bekende uitkomst | Benadering nabestaanden | % bezwaar nabestaan-den indien benaderd |
|---|---|---|---|---|
| Toestemming DR | 229 | 25% | 225 | 12% |
| Bezwaar DR | 158 | 17% | – | – |
| Beslissing nab | 64 | 7% | 608 | 73% |
| Geen registratie | 470 | 51% | | |
| Onbekend | 118 | – | | |
| **Totaal** | **1039** | **100%** | **833** | **56%** |

*Onbekend: bij 118 overledenen werd Donorregister niet geraadpleegd*

Out of 1,039 (100%) potential organ donors, 229 (22%) had consented, 158 (15%) had refused, and 652 (63%) had unknown wishes, among which 64 (6%) had left the decision to their relatives, 471 (45%) were not registered, and in 118 (11%) cases the register was not consulted.

The family was consulted on 833 occasions, both when the deceased had consented (225/833) and when his/her wishes were not known (608/833). The family was not consulted on 206 occasions, either because the deceased had already registered a refusal (158/206) or for other undisclosed reasons (48/206).

In the case of registered consent by the deceased, the family authorized organ retrieval on 198 occasions and opposed a veto on 27 occasions. In the case of unknown preferences from the deceased, the family authorized organ retrieval on 164 occasions and opposed it on 444 occasions.

Table 8

| | | | **Family preferences** | | |
|---|---|---|---|---|---|
| **N=1,039 potential donors** | | | **(a) favorable** | **(b) unfavorable** | **(c) unknown (family not consulted)** |
| | | | *35% (N=362)* | *45% (N=471)* | *20% (N=206)* |
| **Deceased** | **(A) consent** (registered) | 22% (N=229) | *19% (N=198)* | *3% (N=27)* | *0% (N=4)* |
| | **(B) refusal** (registered) | 15% (N=158) | — | *0% (N=0)* | *15% (N=158)* |
| | **(C) unknown preferences*** | 63% (N=652) | *16% (N=164)* | *43% (N=444)* | *4% (N=44)* |

* Decision left to relatives 6% (N=64); Deceased Unregistered 45% (N=471); Register not consulted 11% (N=118)

### Sweden

Source: Nolin, T., Mårdh, C., Karlström, G., & Walther, S. M. (2017). Identifying opportunities to increase organ donation after brain death. An observational study in Sweden 2009–2014. Acta Anaesthesiologica Scandinavica, 61(1), 73–82. https://doi.org/10.1111/aas.12831 [54]

A prospective observational study of all ICU death in Sweden (from Jan. 1, 2009 to Dec. 31, 2014) found 1,575 confirmed potential donors, 240 of which had contraindications to organ donation. Therefore, there was a total of 1,275 eligible organ donors during that 6 years period. Among those 1,275 eligible donors, organ procurement did not proceed because:

- the deceased had expressed their refusal to donation in 176 cases;
- when the deceased's decision was unknown, the family opposed organ recovery in 175 cases;
- when the deceased's decision was unknown, the family could not express a decision in 20 cases (they could not be informed: N=9; they disagreed with one another: N=11).

The resulting 904 eligible donors had either expressed their individual consent or their family authorised recovery:

- the deceased had expressed their consent to donation in 449 cases;
- when the deceased's decision was unknown, the family authorised organ recovery in 455 cases;

Among these 904 eligible donors, the next-of-kin eventually expressed their opposition in 2 additional cases. From the article's description, it seems that these 2 family oppositions should be retracted from the 449 cases where the deceased had consented. Organ recovery was not carried out in 50 additional cases for other reasons (new medical contraindication, no recipient, circulatory collapse, etc.). Eventually, organ recovery was carried out in 852 cases (actual organ donors) out of 904.

During the 6 years period examined, the deceased's preferences and the family's preferences were both unknown (Cc) in 20 cases. Organ recovery did not proceed in these cases. Therefore, during six years (2009-2014), no organs were recovered based on presumed consent. All organs were recovered based on either the deceased's consent or family authorisation.

Table 9

| | | | **Family preferences** | | |
| --- | --- | --- | --- | --- | --- |
| **N=1,275 eligible donors** | | | **(a) favorable** | **(b) unfavorable** | **(c) unknown (family not consulted)** |
| | | | *n/a* | *n/a* | *50% (N=643)* |
| **Deceased's decision** | **(A) consent** | 35% (N=449) | *n/a* | *0% (N=2)* | *35% (N=447)* |
| | **(B) refusal** | n/a | — | *n/a* | *14% (N=176)* |
| | **(C) unknown preferences** | 51% (N=650) | *36% (N=455)* | *14% (N=175)* | *2% (N=20)* |

* Family not informed (N=9); Disagreement among family members (N=11).

### The United Kingdom

Source: NHS, Potential Donor Audit (1 April 2017 - 31 March 2018) [6].

At the end of March 2019, there had been 1,636 potential DBD organ donors (i.e. with no medical contraindications at this stage), and 1,493 requests for authorisation conveyed to their families. The overall consent/authorisation rate was 72.5% (1,082 authorisations over 1,493 family requests):

- When the patient's decision (by any method) was known: 566 consents over 595 requests = 95.13%;
  - 29 families overruled their loved one's known decision (by any method) to be an organ donor.
  - When the patient's decision on ODR and known: 488 consents over 516 requests;
  - Therefore: when the patient's decision was known by other means: 566-488=78 consents over 595-516=79 requests
- When the patient had not expressed a decision or the patient's ODR status was not known: 516 consents over 898 requests = 57.46%
  - However, among the 411 family oppositions, the reason was that the patient previously expressed a wish not to donate on 82 occasions (Table 13.10). Hence, these are 82 cases of known refusal, and unknown ODR status is 898-82=816.

Table 10

| Decision | | Family | | |
|---|---|---|---|---|
| | | Authorisation | Opposition | Total requests |
| Consent | ODR | 488 | 28 | 595 (39.85%) |
| | Other means | 78 | 1 | |
| Refusal | | — | 82 | 82 (5.49%) |
| Unknown | | 516 | 300 | 816 (54.66%) |
| Total | | 1,082 | 411 | 1,493 |

When families are not consulted, there is no data available regarding the deceased´s preferences. However, we can extrapolate the numbers by assuming that there is a similar proportion of consents, refusals, and unknown wishes when families are consulted than when they are not. Therefore, we can complete column c with these numbers.

Table 11

| **N=1636 potential DBD donors** | | | **Family preferences**<br>**(a) favorable** | **(b) unfavorable** | **(c) unknown (family not consulted)** |
|---|---|---|---|---|---|
| | | | *66% (N=1,082)* | *25% (N=411)* | *9% (N=142)* |
| **Deceased** | **(A) consent** (by any means*) | 40%** (N=651) | *35% (N=566)* | *2% (N=29)* | *3%** (N=56)* |
| | **(B) refusal** (oral) | 5%** (N=90) | — | *5% (N=82)* | *0%** (N=8)* |
| | **(C) unknown preferences** (ODR status) | 55%** (N=894) | *32% (N=516)* | *18% (N=300)* | *5%** (N=78)* |

*ODR status (488 authorisations over 516 family approaches) or other means (78 authorisations over 79 family approaches)
** These figures are extrapolated. In 21 cases the patient had registered a decision to donate

#### Wales

Sources:

— [55] Noyes J, McLaughlin L, Morgan K, et al. Short-term impact of introducing a soft opt-out organ donation system in Wales: before and after study. BMJ Open 2019;9:e025159. doi:10.1136/bmjopen-2018-025159

— [6] NHS, Potential Donor Audit (1 April 2017 - 31 March 2018).

During the 15-month period following the implementation of the opt-out (so called “deemed consent”) system, out of 182 deceased adults, 102 had expressed their decision to donate (56%), 34 had expressed their decision to opt-out (18.7%), and 46 had not expressed any preference (25.3%).[1] In all cases, the family was consulted and had the opportunity to either support (123/182) or overrule (13/182) the deceased’s decision[1], and to support (28/182) or oppose (18/182) the recovery of organs when the deceased’s had not expressed any preference (deemed consent).[1] In the UK at large, which includes both opt-in and opt-out systems, circumstances where the family is either untraceable, considered too upset to approach, or divided over the decision were rare (1,6 percent of all eligible donors) between 2017-18.[2] Under those circumstances, organ recovery did not proceed, even in the presence of an explicit consent from the deceased (in opt-in systems) or a deemed consent (in opt-out systems).[2] Wales opt-out default (scenario Cc) was never applied [1].

Table 12

| **N=182 potential donors** | | | **Family** (a) authorises | **Family** (b) opposes | **Family** (c) unknown preferences (family not consulted) |
|---|---|---|---|---|---|
| | | | *64% (N=117)* | *36% (N=65)* | *0% (N=0)* |
| **Deceased** | **(A) consent** | 56% (N=102) | *49% (N=89)* | *7% (N=13)* | *0% (N=0)* |
| | **(B) refusal** | 19% (N=34) | — | *19% (N=34)* | *0% (N=0)* |
| | **(C) unknown preferences** | 25% (N=46) | *15% (N=28)* | *10% (N=18)* | *0% (N=0)* |

1 We assume that family opposition only applies to the deceased’s decision.

#### The United States of America

Source: “National Survey of Organ Donation Attitudes and Practices”, U.S. Department of Health and Human Services, 2019. [Online: https://www.organdonor.gov/sites/default/files/about-dot/files/nsodap-organ-donation-survey-2019.pdf]

The full tables of responses can be found at: https://www.organdonor.gov/sites/default/files/about-dot/files/nsodap-full-response-tables.xlsx

We found some data regarding consent and refusal by the deceased an their family. For instance, data from a study on all eligible death from 2008 to 2013 indicates that, among 52,571 eligible donors, authorisation for donation was based on registration in a donor registry in 7,562 cases (14%), and it was obtained from family or next-of-kin (with or without prior registration in a donor registry) in 30,870 cases (59%) [56]. See also [48,49]. However, we couldn’t find detailed enough actual data from the US to fill the scenarios table.

Instead, we will consider an hypothetical situation based on stated attitudes towards donation, under the assumption that these attitudes reflect actual behaviour, even though we know that this not necessarily the case. For that reason, we do not include the US in Table 3 of the manuscript.

A US national survey of public attitudes on a sample of 10.000 individuals found half (49,9%) of respondents had signed up as donors (N=4,990). Of those who were not signed up as donors (46.2%, N=4,620), half said they wished to donate their organs after death (50.3%, N=2,324), less than half said they wished not to donate (46.7%, N=2,158), and 3% (N=139) didn’t answer. Among those who signed up as donors, 68.9% (N=2,438) discussed their wish with a family member, and 27.2% didn’t discuss it. Among those who wished not to donate, 23.5% (N=507) discussed their wish not to donate with their family while 59.6% didn’t discuss it. We don’t know about family discussions of those who wished to be donors but didn’t sign up. However, to the question “Has any member of your family told you about his or her wish to donate or not to donate his or her organs after death?”, 45,6% answered “yes” and 54,4% answered “no”. Considering that this question includes both willingness and unwillingness to donate, and considering that individuals tend to have more than one family member, we will suppose that among those who wished to donate but didn’t sign up (N=2,324), only 30% (N=697) discussed their wish with their family.

Therefore, regarding the deceased’s preferences:

- Known consent to donate: 5,687
  - Signed up as donors: 4,990
  - Didn’t sign up but discussed it with family: 697
- Known refusal to donate: 507
  - Discussed it with family: 507
- Unknown wishes: 3,806
  - Didn’t answer about their wishes: 139
  - All those who didn’t sign up nor discussed their wishes: 3,667

About willingness to donate family member’s organs: 88,3% would donate a family member’ organs if their wishes to donate were know and 11,7% would not; while 68,8% say they would donate a family member’s organs if their wishes were unknown and 31,2% would not. These responses allow us to fill the table:

- Deceased’s known consent and family authorisation: 88,3% of 5,687 = 5,022
- Deceased’s known consent and family opposition: 11,7% of 5,687 = 665
- Deceased’s known consent and unknown family decision: 5,687-5,022-665=0

    - However, we can’t rule out the possibility that the deceased had no family, that the family could not be contacted on time, or that they didn’t agree between each other.
- Deceased’s unknown wishes and family authorisation: 68,8% of 3,806 = 2,619
- Deceased’s unknown wishes and family opposition: 31,2% of 3,806 = 1,187
- Deceased’s unknown wishes and family unknown wishes: 3,806-2,619-1187=0
- Decesead’s known refusal and family authorisation: although some family members would consider overruling their loved one refusal (11% according to Sellers et al. 2018), this option is currently not allowed by consent systems. For that reason, we will consider it as nil. Anyway, there is no data in this survey about that option.
- Deceased’s known refusal and family opposition: no data available
- Deceased’s known refusal and family unknown wishes: all individual refusal cases = 507

Table 13

| | | | Family | | |
|---|---|---|---|---|---|
| **N=10,000 respondents** | | | **(a) authorises** | **(b) opposes** | **(c) unknown preferences** |
| **Deceased** | **(A) consent** | 57% (N=5,687) | *50% (N=5,022)* | *7% (N=665)* | *0* |
| | **(B) refusal** | 5% (N=507) | — | *n/a* | *5% (N=507)* |
| | **(C) unknown preferences** | 38% (N=3,806) | *26% (N=2,619)* | *12% (N=1,187)* | *0* |

Our result for scenario Ab (7%) is consistent with a survey of all 58 Organ Procurement Organizations (OPO) in the US that estimated that the frequency of family objecting to organ donation in cases of registered donors was <10% [57]. However, following the First Person Authorization policy, most OPOs would ignore the family opposition and proceed to organ donation anyway: “Over the past 5 years, 65% of OPOs have participated in procuring organs even when the next-of-kin and the deceased have differing opinions regarding organ donation [and the remaining 35% of OPOs have not participated in organ procurement from a registered organ donor in the setting of family objection]. Nearly 80% of OPOs reported that this situation occurred one to five times, whereas the remaining 20% reported more than five occasions during that period.” [57]. Therefore, the figure of 7% in scenario Ab may correspond to family oppositions but not to actual outcomes (organs not being procured in that circumstance).

To include the US in figure 1 as an hypothesis, we need to tweak the figures from attitudes to reflect more realistic data about actual outcomes. This is the likelihood of procuring organs in different scenarios according to the OPOs survey [57]:

**Table 1:** Comparison of the likelihood of procuring organs between the FPA-compliant and FPA-noncompliant OPOs in the settings where the deceased and the family's wishes differ or are not known

| Scenario | Deceased | Family | % FPA NC OPOs likely to procure | % FPA C OPOs likely to procure | p-Values |
|---|---|---|---|---|---|
| A | Registered | Objects to donation | 45% (5/11) | 100% (42/42) | <0.001 |
| B | Registered | Unavailable | 100% (11/11) | 98% (41/42) | NS |
| C | Registered | Unable to reach decision | 82% (9/11) | 100% (42/42) | 0.04 |
| D | Object to donate | Unavailable or unable to reach decision | 30% (3/10) | 5% (2/42) | 0.043 |
| E | Wish unknown | Unavailable or unable to reach decision | 45% (5/11) | 29% (12/42) | NS |
| F | Wish unknown | Would like to donate | 100% (11/11) | 100% (42/42) | – |
| G | Wish unknown | Objects to donation | 0% (0/11) | 2% (1/42) | NS |

FPA C, first person authorization compliant Organ Procurement Organizations (OPOs); FPA NC, first person authorization noncompliant OPOs.

*American Journal of Transplantation* 2014; 14: 172–177 **175**

Likelihood of procuring organs in these scenarios is:

A (Ab): 47 OPOs (5+42) out of 53 /(11+42) = 88,6%

B (Ac): 52 out of 53 = 98,1%

C (Ac): 51 out of 53 = 96,2%

D (Bc): 5 out of 52 = 9,6% (sic! This means that OPOs may procure organs despite the deceased's refusal and without an authorization from the family)

E (Cc): 17 out of 53 = 32,1%

F (Ca): 53 out of 53 = 100%

G (Cb): 1 out of 53 = 1,9%

In addition, we will consider that unknown family wishes in scenarios Ac and Cc are 2% (N=200) each. These figures are retracted proportionally from the other scenarios:

Table 14

| | | | **Family** | | |
|---|---|---|---|---|---|
| | **N=10,000 respondents** | | **(a) authorises** | **(b) opposes** | **(c) unknown preferences** |
| **Deceased** | **(A) consent** | 57% (N=5,687) | *48% (N=4,845)* | *6% (N=642)* | *2% (N=200)* |
| | **(B) refusal** | 5% (N=507) | — | *n/a* | *5% (N=507)* |
| | **(C) unknown preferences** | 38% (N=3,806) | *24% (N=2,481)* | *11% (N=1,125)* | *2% (N=200)* |

Therefore, the likelihood of procuring organs in the current system (opt-in) and considering the family wishes is:

— When the deceased's consent is known (Aa: 4,845; Ab: 642*88,6%=569; Ac: 200*96,2%=192): 5,606

— When the deceased's refusal is known (Bc: 507*9,6%=49): 49

— When the deceased's wishes are unknown (Ca: 2,481*100%=2,481; Cb: 1,125*1,9%=21; Cc: 200*32,1%=64): 2,566

— Total: 8,221 out of 10,000 = 82%

If in the current system (opt-in) the family wishes were ignored:

— When the deceased's consent is known: 5,687

— When the deceased's refusal is known: 0

— When the deceased's wishes are unknown: 0

— Total: 5,687 out of 10,000 = 57%

If in an opt-out system and the family wishes were considered (as they are now): same as in opt-in but adding the 200 cases of scenario Cc. Total: 8,221+200=8,421 –> 84%

If in an opt-out system and the family wishes were ignored:

— When the deceased's consent is known: 5,687

— When the deceased's refusal is known: 0

— When the deceased's wishes are unknown: 3,806

— Total: 9,493 out of 10,000 = 95%

Table 15: Data used to draw Fig. 1 of the article

| | Deceased's wishes alone | | Deceased's and family wishes | |
|---|---|---|---|---|
| Country | Opt-in | Opt-out | Opt-in | Opt-out |
| Denmark | 6 | 84 | 35 | 37 |
| Germany | 32 | 91 | 74 | 76 |
| The Netherlands | 22 | 85 | 35 | 39 |
| Sweden | 35 | 86 | 71 | 72 |
| United Kingdom | 40 | 94 | 70 | 74 |
| Wales | 56 | 81 | 64 | 64 |
| United States | 57 | 95 | 82 | 84 |

Table 15bis

| | Opt-in | | | Opt-out | | |
|---|---|---|---|---|---|---|
| Country | Deceased alone | Deceased + family | Variation | Deceased alone | Deceased + family | Variation |
| Denmark | 6 | 35 | 29 | 84 | 37 | -47 |
| Germany | 32 | 74 | 42 | 91 | 76 | -25 |
| The Netherlands | 22 | 35 | 13 | 85 | 39 | -46 |
| Sweden | 35 | 71 | 36 | 86 | 72 | -14 |
| United Kingdom | 40 | 70 | 30 | 94 | 74 | -20 |
| Wales | 56 | 64 | 8 | 81 | 64 | -17 |
| United States | 57 | 82 | 25 | 95 | 84 | -11 |

3. European Commission. Special Eurobarometer 333a: Organ donation and transplantation. Brussels; 2010.

4. CBS. Nearly half of Dutch population included in Donor Register. In: Statistics Netherlands [Internet]. 17 Jun 2020 [cited 6 Aug 2020]. Available: https://www.cbs.nl/en-gb/news/2020/25/nearly-half-of-dutch-population-included-in-donor-register

5. Caille-Brillet A-L, Zimmering R, Thaiss HM. Bericht zur Repräsentativstudie 2018 „Wissen, Einstellung und Verhalten der Allgemeinbevölkerung zur Organ- und Gewebespende“. Köln: BZgA-Forschungsbericht; 2019.

6. NHS. Transplant activity report. In: NHS Organ Donation [Internet]. 2019 [cited 6 Aug 2020]. Available: /helping-you-to-decide/about-organ-donation/statistics-about-organ-donation/transplant-activity-report/

12. Molina-Pérez A, Delgado J, Rodríguez-Arias D. Defining Consent: Autonomy and the Role of the Family. In: Hansen SL, Schicktanz S, editors. Ethical Challenges of Organ Transplantation: Current Debates and International Perspectives. Transcript Verlag; 2021.

13. Rosenblum AM, Horvat LD, Siminoff LA, Prakash V, Beitel J, Garg AX. The authority of next-of-kin in explicit and presumed consent systems for deceased organ donation: an analysis of 54 nations. Nephrol Dial Transplant. 2012;27: 2533–2546. doi:10.1093/ndt/gfr619

14. Bilgel F. The impact of presumed consent laws and institutions on deceased organ donation. Eur J Health Econ. 2012;13: 29–38. doi:10.1007/s10198-010-0277-8

15. DSO. Jahresbericht Organspende und Transplantation in Deutschland 2018. Frankfurt/Main: Deutsche Stiftung Organtransplantation; 2019.

16. Cho W-H. Organ donation in Korea in 2018 and an introduction of the Korea national organ donation system. Korean J Transplant. 2019;33: 83. doi:10.4285/jkstn.2019.33.4.83

17. INCUCAI. Procuración y trasplante de órganos, tejidos y células en Argentina. Memoria 2018. 2019.

18. INCUCAI. Procuración y trasplante de órganos, tejidos y células en Argentina. Memoria 2019. 2020.

19. Melville A, Kolt G, Anderson D, Mitropoulos J, Pilcher D. Admission to Intensive Care for Palliative Care or Potential Organ Donation: Demographics, Circumstances, Outcomes, and Resource Use. Critical Care Medicine. 2017;45: e1050–e1059. doi:10.1097/CCM.0000000000002655

20. SBT. Rapport annuel 2019 - Section belge des coordinateurs de transplantation - SBT. Service Public Fédéral; 2016 Apr.

21. Dell Agnolo CM, de Freitas RA, Toffolo VJO, de Oliveira MLF, de Almeida DF, Carvalho MDB, et al. Causes of Organ Donation Failure in Brazil. Transplantation Proceedings. 2012;44: 2280–2282. doi:10.1016/j.transproceed.2012.07.133

22. Rosário EN do, Pinho LG de, Oselame GB, Neves EB. Recusa familiar diante de um potencial doador de órgãos. Cad saúde colet. 2013;21: 260–266. doi:10.1590/S1414-462X2013000300005

23. Marinho BBO, Santos ATF, Figueredo AS, Cortez LSAB, Viana MCA, Santos GM, et al. Challenges of Organ Donation: Potential Donors for Transplantation in an Area of Brazil's Northeast. Transplantation Proceedings. 2018;50: 698–701. doi:10.1016/j.transproceed.2018.02.055

24. Corporación del Trasplante. Memoria Anual 2008. Corporación del Trasplante, Chile; 2009. Available: https://studylib.es/doc/8240405/memoria-2009---corporaci%C3%B3n-del-trasplante

25. Instituto Nacional de Salud. Informe anual Red de Donación y Trasplantes. Instituto Nacional de Salud, Colombia; Available: https://www.ins.gov.co/Direcciones/RedesSaludPublica/DonacionOrganosYTejidos/Paginas/default.aspx

35. Health Service Executive. Audit of Potential Organ Donors, Republic of Ireland. Government of the Republic of Ireland, Health Service Executive; 2009.

45. Immer FF, Comité National du Don d'Organes. Variation in Organ Donation Rates in Switzerland: Prospective Cohort Study of Potential Donors (SwissPOD). Swisstransplant; 2014.
